# The impact and experience of a culturally adapted brief intervention for harmful alcohol use in an emergency care setting (PPKAY: Punguza Pombe Kwa Afya Yako): A case series

**DOI:** 10.1101/2025.10.09.25337396

**Authors:** Kim Madundo, Anaelderrick Kimaro, Ashley Phillips, Linda Minja, Msafiri Pesambili, João Vitor Perez de Souza, Alice Andongolile, João Ricardo Nickenig Vissoci, Blandina T. Mmbaga, Catherine Staton

## Abstract

**Background:** Harmful alcohol use is widespread in Tanzania, yet specialized resources to address this remain limited. This case series explores outcomes of three individuals who received a culturally-adapted brief intervention to reduce harmful alcohol use in an emergency department in Kilimanjaro, Tanzania.

**Methods:** The PPKAY intervention is a 15-minute nurse-delivered motivational interview for adults with acute (<□24□h) injuries, with an Alcohol Use Disorder Identification Test (AUDIT) of 8 or above or self-reported drinking alcohol within 6 hours of injury. The primary outcome was participants’ experience of the intervention. Secondary outcomes included number of binge drinking days in the prior month; depression, measured using the Patient Health Questionnaire-9 (PHQ-9); and alcohol-related consequences, measured by the Drinker Inventory of Consequences (DrInC). All outcomes were reported from baseline and at 3-month timepoints. Experience with the intervention was assessed with open-ended questions at a three-month post-intervention survey.

**Results:** Participants reported fewer binge drinking days, improved self-efficacy in managing triggers, reduced alcohol-related consequences, remission of depression, and strengthened interpersonal relationships. These were corroborated by significantly lower AUDIT, PHQ-9, and DrInC scores. One participant was not able to meet his drinking reduction target, accompanied by an avoidance of discussing interpersonal consequences of alcohol.

**Conclusions:** Our findings underscore the importance of integrated and targeted early screening and intervention in resource-limited emergency care settings. The narratives emphasize how a brief negotiational intervention can equip individuals with practical strategies to minimize harmful alcohol consumption. Further research is recommended to validate these outcomes in larger cohorts and diverse settings.

## Background

Harmful alcohol use poses a significant public health issue worldwide, contributing to a considerable burden of morbidity and mortality. According to the World Health Organization (WHO), alcohol consumption accounts for nearly 5% of all global deaths (Hammer et al., 2018; World Health Organization, 2024a). This burden is especially high in low- and middle-income countries (LMICs), where the harmful effects of alcohol are exacerbated by minimal enforcement of regulations, low awareness of alcohol-related harms (Rehm et al., 2009; Rehm & Poznyak, 2015), and the integral role of alcohol in socio-cultural practices (Madundo et al., 2024).

Tanzania faces disproportionately high levels of these harms, where the risk of alcohol use disorder (AUD) is six times the African average, and the rates of binge drinking are twice the average for Africa (Hammer et al., 2018). Additional harms fall under physical, social responsibility, interpersonal, intrapersonal, and impulse control domains (Zhao et al., 2018); these impacts include acute injuries, unplanned pregnancies, violence, non-communicable diseases, and co-morbid mental disorders (Ferreira-Borges et al., 2017; Staton et al., 2018). In the Kilimanjaro region of Tanzania, alcohol use is deeply ingrained in cultural and social contexts, presenting further challenges to interventions (Madundo et al., 2024; Massawe et al., 2022). Studies from this region revealed that 30% of acute injury patients test positive for alcohol upon arrival and that alcohol consumption increases the risk of injury by 5.7 times (Staton et al., 2018). Further, more than 80% of the alcohol consumed comes from unregulated local brews, which further complicates efforts to mitigate alcohol-related harm (Ibitoye et al., 2019; Madundo et al., 2024).

Despite the clear need for targeted interventions, Tanzania faces a severe shortage of mental health resources and evidence-based programs to address harmful alcohol use (Knettel et al., 2023; Mushi, Francis, et al., 2022). Literature reveals that approximately one mental health professional is available for more than one million Tanzanians (Knettel et al., 2023), in a highly fragmented health system lacking transition from hospitals to scarce community-based care (Faro et al., 2023). A potential solution to these challenges is brief negotiation interventions (BIs) for alcohol use; cost-effective, less resource-intensive, and effective in reducing alcohol-related harms. However, BIs have been developed in Western contexts and require cultural adaptation to be effective in African settings (Ghosh et al., 2022; Zimmerman et al., 2021). Culturally adapted BIs could provide a promising approach to mitigating the impact of harmful alcohol use in emergency department (ED) settings, where patients may be particularly receptive to behaviour change messages (Zimmerman et al., 2021). This approach aligns with the WHO’s SAFER initiative to increase access to alcohol screening, brief interventions, and treatment as a strategy to minimize related harms (World Health Organization, 2019, 2024b).

This study presents a series of three cases describing the impact of a culturally adapted BI for the reduction of alcohol use and its associated harms (Zhao et al., 2018), and the participants’ experiences of this intervention conducted in an ED setting. The three patients had no prior experience of counselling or BIs.

## Methods

This current study is part of a randomized clinical trial evaluating the effectiveness of BI at reducing hazardous alcohol drinking among patients with acute injuries. The intervention is named PPKAY, abbreviated for ‘Punguza Pombe Kwa Afya Yako’, Swahili for ‘reduce alcohol use for your health’). The full trial protocol is registered at ClinicalTrials.gov NCT04535011, with Protocol no. Pro000103724. The primary outcome for the overall trial was the number of binge-drinking events in the past month, and this study is described in further detail elsewhere (Staton et al., 2022).

### Study Setting

The parent study was conducted in the emergency department (ED) at Kilimanjaro Christian Medical Centre (KCMC), the third-largest hospital in Tanzania and a tertiary-level academic facility in an urban resource-limited setting. The ED serves a catchment area of 15 million people and receives approximately 24,000 patients annually. Of this number, 17% present with acute injuries per year. For every 10 patients with an acute injury at the ED, three are intoxicated with alcohol (Staton et al., 2018).

### Clinical Trial Procedures

The PPKAY intervention, a nurse-delivered motivational interview (MI), was designed to reduce hazardous alcohol use among adults with acute (<□24□h) injuries (Staton et al., 2022; Zimmerman et al., 2021). Upon arrival at the ED and before the intervention, patients were screened by a nurse using the Swahili-validated Alcohol Use Disorder Identification Test (AUDIT) and tested by breathalyzer.

Inclusion criteria were the presence of an acute injury, AUDIT score of ≥8, age 18 or older, and/or positive alcohol breathalyzer test (>□0.0□g/d□). Exclusion criteria were inability to speak Swahili and being clinically intoxicated based on history and physical examination.

Patients who met the selection criteria were invited to participate in the study and referred to a study nurse who asked permission to discuss recent alcohol use further. Written informed consent was obtained by research assistants (RAs), who also explained the study’s aims and the randomization process. Concerns about confidentiality were addressed during the consenting process. The RAs then conducted a baseline survey comprised of the AUDIT, Patient Health Questionnaire-9 (PHQ-9), the Drinker Inventory of Consequences (DrInC), and questions on drinking patterns over the past month.

The intervention took place in a private area of the ED to ensure confidentiality and comfort during the session. The duration of the intervention was strictly maintained, lasting approximately 30 minutes for each participant, from consent to the end of the BI.

Participants randomized to the BI study arm met with a nurse for the 15-minute intervention in a four-step process: (1) raise the subject of alcohol, (2) provide feedback, (3) enhance motivation, and (4) negotiate and advise. The nurse asked permission to audio-record the session and engaged the participant in a non-judgmental discussion about alcohol use, defined standard drink units, and explained the AUDIT tool as a means to determine hazardous levels of alcohol consumption. The nurse then asked the participant to interpret their alcohol use based on their AUDIT score, prompted responses regarding consequences of drinking, and reflected on these connections. The nurse employed a brief MI approach to identify stage of behaviour change and readiness to change on a scale from 0-100%. Resistance to change was negotiated to guide and enhance motivation, for example: “You reported 80% readiness to change, why didn’t you choose a lower number?” This approach placed emphasis on the positivity, and preparedness for change (Staton et al., 2022; Zimmerman et al., 2021).

The nurse and participant then collaboratively negotiated and set achievable short-term goals aimed at reducing alcohol-related harm. Participants were praised for their solutions and given supportive advice on their chosen actions. Information about follow-up care and support services was shared with participants, such as mental health services at KCMC and rehabilitation services in Kilimanjaro region. Participants were reminded that information from this session would be used to tailor text messages. Lastly, the nurse summarized the patients’ statements and words of change, emphasized strengths, reflected on agreements, and asked for any further questions. After the intervention, participants were assessed on their immediate response to the session, and follow-up data were collected at a three-month follow-up and up to two-year follow-up by survey (Staton et al., 2022; Zimmerman et al., 2021).

### Current Study Procedures

This case series focuses on three participants who received the PPKAY BI with no modifications to the protocol. The unique cases were purposively selected from the intervention arm of the clinical trial based on reviews of audio recordings of BI sessions and case presentations made during weekly clinical supervision sessions with two research coordinators (AP and LM) and a practicing psychiatrist (KM).

The cases describe individuals who presented unique challenges related to harmful alcohol use, their mental health, and other relevant life experiences. All three cases highlight the varying levels of success of the BI and describe individuals with no prior experience of counselling or mental health services.

### Measures and Outcomes

The primary outcome for this sub-study was the experience of the BI. Secondary outcomes were based on quantitative measures: number of binge-drinking events in the past month, AUDIT scores (≥8); depression, measured using the Patient Health Questionnaire-9 (PHQ-9) with a cut-off score of 9; and perceived alcohol-related consequences measured by the Drinker Inventory of Consequences (DrInC). These measures have been validated for use in Tanzania through independent forward- and back-translation processes, cognitive interviews by the translators and the study team to resolve discrepancies in the translated versions, and to ensure cultural appropriateness (Smith Fawzi et al., 2019; Vissoci et al., 2018; Zhao et al., 2018).

We assessed the participants’ experiences of the intervention by asking about the acceptability and perceived usefulness of the BI. These data were collected as prompted qualitative feedback at the end of the three-months post-intervention survey, however, were not described in the main trial protocol publication (Staton et al., 2022). There were no adverse or unanticipated events to report.

### Analysis

Quantitative outcomes were assessed at baseline and compared at 3-months post-intervention. Qualitative feedback was entered as text in a spreadsheet by a RA at the follow-up survey. KM, AK, AP, and MP used a deductive process to analyze the quotes. A coding framework was developed post-survey using the DrInC domains, and refined during analysis. The quotes were coded to the following fields: physical, social, interpersonal, intrapersonal, and impulse control. Key themes were identified by KM and AK, and discussed by the larger team. Coherent narratives were thereafter created by aligning themes from the organized quotes.

We present three case reports with both quantitative and qualitative findings. Integration of these mixed methods was set at the design stage of the current study (considering both quantified scores and qualitative feedback in the study aims), data collection stage (collecting both quantitative and qualitative data), and in the discussion (using qualitative feedback to further explain and support the quantitative findings). We triangulated these findings at the investigator level: the first author viewed and analyzed the data separately from other team analysis members to increase the chance of uncovering errors and minimize investigator bias. Thereafter all authors reviewed and edited the work for accuracy.

### Ethical Considerations

This study was approved by the ethics review committees at Duke (Pro000106116), Kilimanjaro Christian Medical Centre (Cert. #2486) and Tanzania’s National Institute of Medical Research (HQ/R.8a/Vol.IX, 3425). All participants provided written informed consent before participation in the study.

The intervention was designed to minimize any risk or discomfort for participants. The cases have been deidentified and details, including names and age, have been altered to ensure participant confidentiality without affecting the quality and accuracy of the case reporting. Audio recordings, spreadsheets and analysis documents were stored on a password-protected server only accessible to the research team, and recordings were erased from voice recorders immediately after upload.

If any participant reported significant distress related to alcohol use or related injuries, they were offered referrals to appropriate follow-up care or mental health services. Emergency contact information was provided to all participants for crisis intervention if needed.

## Case presentations

### The case of Massawe

Massawe was a man in his late 20’s, a self-employed farmer, with secondary school level of education. He reported an estimated monthly income of $75 (200,000 Tanzanian shillings), whereby the minimum monthly wage for public officials is $198 (Okafor, 2025).

Massawe was referred from a district-level hospital, presenting with an accidental acute injury after falling from a tree. He was referred due to diagnostic and treatment challenges in primary care.

At baseline, Massawe reported eight binge drinking events within the past month. He scored 24 on the AUDIT, indicating high likelihood of severe alcohol use disorder. His PHQ9 score indicated no depression, however, the DrInC score was suggestive of moderate-level consequences due to drinking including interpersonal, physical and social aspects (Zhao et al., 2018).

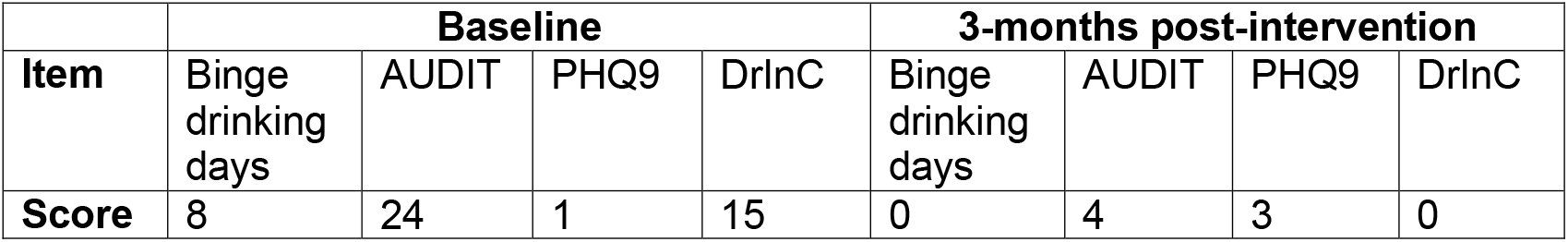

Massawe opened up quickly during the 15-minute nurse-guided session. He was motivated to cut down on his drinking for two: reasons; primarily, to save money, and also to live an overall healthier life. He recognized the negative influence alcohol had on his work and personal life, and vowed to avoid situations where he would be pressured to consume alcohol. He believed that spiritual support would also be crucial during this transition.

> *“I need to keep myself busy with farming, and to avoid the friends who drink a lot. I can also seek help from my pastor”*.

He desired to quit alcohol completely because he noticed that people expressed judgment and mistrust related to his alcohol use.

> *“Everybody said I got injured because I was drunk. They don’t believe in me. I feel like it is ruining my reputation*.*”*

Massawe was admitted to KCMC’s orthopedic surgery ward for urgent surgical intervention. He was later discharged with serious disability requiring assistance with self-care, toileting, and locomotion.

3 months post-intervention, Massawe reported remarkable benefits from the BNI, the most important to him being a significant reduction in alcohol use. Although still dependent on family for movement and self-care, he felt overall healthier, and felt proud of his accomplishments. While cutting down on alcohol he also engaged in mental health services for additional support.

> *“I went Dar es salaam to see a psychologist. We discussed a lot of things which helped me to improve my health*.*”*

This was supported by Massawe’s 3-month post intervention scores, which revealed objective reductions of all scores related to alcohol harms. Massawe had zero binge drinking events within the past month, his AUDIT score was below the cut-off of eight, and his perceived recent consequences of drinking were also resolved. Although the PHQ9 score was slightly elevated, it was still suggestive of no or minimal depression.

When asked about the acceptability of the BNI, Massawe reported resonating with the topics discussed in the intervention. He praised the study staff their competence and the intervention for its relevance.

> *“This [intervention] is so useful. I decided to follow the advice from the [study] nurse because she had important ideas. I listened to her and things went well*.*”*

### The case of Kimaro

Kimaro was a man in his mid-30’s, a self-employed businessman with secondary school level of education. Part of his business included selling alcoholic beverages, where he earned $110 per month.

Kimaro was referred to KCMC’s ED from a regional-level hospital, presenting with accidental injuries from a fall while working in construction. A referral was initiated due to limited treatment capacity for acute injuries at the regional level.

Upon initial presentation, Kimaro estimated 12 binge drinking events within the month prior to injury. His AUDIT score of 28 was further indicative of clinically significant alcohol related harms. Kimaro perceived a moderate level of consequences on his interpersonal and intrapersonal functioning, and the PHQ9 score of 24 suggested a severe level of depressive symptoms, congruent with the AUDIT and DrInC scores.

|  | **Baseline** |  |  |  | **3-months post-intervention** |  |  |  |
| --- | --- | --- | --- | --- | --- | --- | --- | --- |
| **Item** | Binge drinking days | AUDIT | PHQ9 | DrInC | Binge drinking days | AUDIT | PHQ9 | DrInC |
| **Score** | 12 | 28 | 24 | 16 | 0 | 18 | 4 | 5 |

Despite the motivational approach, Kimaro was cautious in his expectations to cut down on alcohol use. He predicted he would reduce the number of drinks consumed per sitting from eight to five, which would still qualify as a binge drinking event. Kimaro’s cautiousness may be explained by his regular income from selling alcohol, however, he was optimistic that he could terminate this business.

After the 15-minute BNI he was motivated to avoid social situations where heavy drinking was normalized, and the risks that followed.

> *“I need to avoid the friends who drink alcohol so much, which means I need to stop doing this business of selling beer. And I need to stop driving when I’m drunk”*

Kimaro reported the most significant benefit from the BNI was finally accepting that he had problematic drinking behaviours. His injuries were a consequence too difficult to ignore, and the intervention helped him enact the necessary decisions.

> *“I just had to accept that there was a problem. I mostly needed to reduce the amount of alcohol I was using, but I still changed my job because my foot is still not good*.*”*

Kimaro further reflected on other areas of his life which he had been ignoring, such as financial stability and physical health.

> *“Nowadays I have money in my pocket at the end of the day. I can control my income and I have savings. I listened to the nurse when she told me that alcohol was bad for my health, and I now see the difference”*

He was admitted to KCMC’s orthopedic surgery ward where he received urgent surgical care. Kimaro was later discharged with mild discomfort (pain 2/10), although requiring assistance with hygiene, walking, and climbing stairs.

The 3-month post intervention scores indicated that Kimaro had exceeded his expectations and reduced the number of binge drinking events to zero. He had also achieved significant independence in self-care and mobility. Similarly, his most recent depression scores and perceived level of consequences related to alcohol were minimal. Although his AUDIT score (18) was still suggestive of severe alcohol use disorder, this is a reflection of the AUDIT’s focus on broader ‘past year’ alcohol use, not specifically binge drinking.

Kimaro reported overall acceptance of the BNI without any complaints.

### The case of Lyimo

Lyimo was a man in his early 50’s employed as a seasonal skilled manual labourer. He had completed a primary level of education. He declined to disclose his monthly income.

Lyimo was referred from a regional-level health facility and presented with an injury resulting from a blunt force injury inflicted by another person. Similar to Kimaro, Lyimo’s referral was initiation due to limited diagnostic and treatment capacity. Both Lyimo and the offender were under the influence of alcohol at the time of injury.

His history of alcohol use included 10 binge drinking events within the month before the current acute injury. Similarly, he had a high AUDIT score of 19, indicating likelihood of severe alcohol use disorder, and very high perceived level of intra- and interpersonal, social, and physical consequences due to his alcohol use (Zhao et al., 2018). Despite these scores, he presented with mild probable depressive symptoms.

|  | Baseline |  |  |  | 3-months post-intervention |  |  |  |
| --- | --- | --- | --- | --- | --- | --- | --- | --- |
| Item | Binge drinking days | AUDIT | PHQ9 | DrInC | Binge drinking days | AUDIT | PHQ9 | DrInC |
| Score | 10 | 19 | 8 | 45 | 8 | 9 | 5 | 12 |

Lyimo referred to feelings of guilt around his heavy drinking, and how this was affecting his wife and children, and the connection with his mother. Accordingly, he was motivated to quit drinking completely so as to be a better parent and husband. During the 15-minute session, he expressed high optimism about his ability to better manage his finances, to attend mental health clinic for additional support, and to avoid feeling pressured to drink by his peers.

*“I want to stop drinking completely. I want to be a better father. I will attend clinic and avoid all people who drink at the bar. I will save money to help my family*.*”*

Lyimo was thereafter admitted to KCMC’s orthopedic surgery ward where he received urgent surgical care. He was later discharged with moderate pain (4/10) and functionally, required minimal assistance with his daily activities.

At the follow-up survey, Lyimo reported that demonstrating effort to cut down on drinking had improved his relationships with his mother. He had achieved almost total independence in self-care and mobility. He also realized the negative impact of heavy alcohol use on his mental state.

> *“[After the injury] I was told by many people that I was stressed and depressed. They told me to rest and eat healthy. My mother always reminded me of the benefits of reducing my alcohol use*.*”*

Although Lyimo perceived increased social support, particularly from peers and his mother, he was not able to meet his goals in quitting alcohol at 3-months post intervention. During this survey, he reported eight binge drinking events within the past month and an elevated AUDIT score of 9, both indicative of likely hazardous alcohol use. Lyimo declined to comment directly on the tension with his wife and children, however, reported a significant reduction in the perceived level of interpersonal and intrapersonal functioning.

Similar to the two other participants, Lyimo was grateful for the intervention and did not report any negative feedback.

## Discussion

This PPKAY intervention was delivered by trained nurses within 15 minutes, in a culturally sensitive and resource-constrained environment, and aimed to reduce alcohol consumption and related harms. The findings highlight the potential benefits of such interventions in a LMIC setting, where alcohol-related harm is a critical public health challenge that is largely unaddressed. Here we discuss the impact of the intervention guided by the patients’ experiences of this BI.

### Impact of the Intervention

The BI demonstrated varying levels of success across the three cases, with each participant showing some degree of reduction in alcohol consumption and related harms. All participants reported a decrease in binge drinking days from baseline to the 3-month follow-up timepoint, as well as lower intensity in the recent perceived consequences due to alcohol, and a remarkable decrease in level of depressive symptoms.

The shift across these parameters within a 3-month period, and initiated by a 15-minute nurse-delivered intervention suggests the undeniable impact a culturally adapted BI, and its effectiveness in a resource-limited setting. This is in contrast to some global literature that shows minimal impact of BIs on binge drinking events or drinking intensity (Kaner et al., 2018), however, aligns with findings from studies that reveal larger effect sizes with nurse-delivered BIs (Platt et al., 2016). It is possible that the current BI has shown more promising impact on participants because of their associated acute injuries. Previous literature has revealed a stronger inclination to follow-through on behaviour change and lifestyle modifications when individuals are faced with a critical event (Boudreaux et al., 2012; Van Den Ende et al., 2021). Another possibility is the physical inability to access alcohol during the three months post-injury because of limited mobility. However, this is only true for Massawe as the two other participants had achieved significant independence in self-care and locomotion, indicating that they could consume alcohol if they wished to.

The three cases highlight the importance of addressing not only alcohol consumption but also the broader sociocultural factors that contribute to alcohol misuse. The Kilimanjaro region is known for relatively high consumption of alcohol (Massawe et al., 2022), and for largely unregulated sources of alcohol such as local brew (Madundo et al., 2024; Mitsunaga & Larsen, 2008). For BIs to be effective in a context such as this one, it is paramount to acknowledge the sociocultural aspects of alcohol use, which include production of and selling alcoholic beverages as a source of income (Madundo et al., 2024). The MI component thereby plays a role in holistically weighing the benefits and risks of engaging in alcohol consumption (Smedslund et al., 2011). Previous literature has highlighted the ineffectiveness of interventions that transfer intervention models and ignore contextual differences (Marsiglia & Booth, 2015; Satre et al., 2015).

The qualitative and quantitative findings from the three cases complement each other. Particularly, the participant quotes add depth to our understanding of changes in symptom scores and functional status from baseline to the three-months post-intervention timepoint. Although Massawe demonstrated significant reduction of AUDIT and DrInC scores, he had a slight increase in depressive symptom scores over the three months. This could be explained by his long-term dependence on family for self-care and mobility. A similar theory would explain universal reduction across the AUDIT, DrInC and PHQ-9 scores who had over time achieved significant improvement in his physical and intrapersonal functioning. Previous literature supports this relationship between post-injury quality of life, acquisition of basic needs and depression (Broton et al., 2022; Shin et al., 2012).

In contrast, Lyimo’s case exemplifies the challenges of achieving cessation of alcohol use. While he was able to reduce his binge-drinking by two events per month and achieve lower perceived consequences of drinking, he did not meet his goals. His avoidance of exploring interpersonal functioning suggests deeper challenges that could negatively impact his ability to sustainably reduce his alcohol consumption (Moos & Moos, 2006). This case demonstrates the complexity of substance use disorders; individuals may express enthusiasm and confidence in cutting down on substance use, yet relapses are the rule rather than the exception (Nguyen et al., 2020). Individuals affected by alcohol use disorder often need longer-term support to sustain behaviour change (Nguyen et al., 2020; Smedslund et al., 2011). All participants who received the PPKAY BI were referred to mental health services for continued usual care, however, the services available at KCMC and in Kilimanjaro lack the sustainable controlled environment support and specialized rehabilitation facilities, and may be less effective in eliciting sustained recovery (Substance Abuse and Mental Health Services Administration (SAMHSA), 1997; World Health Organization, 2021). Additionally, there is high fragmentation of mental health systems and services in Tanzania, with little to no transition between acute care and recovery in the community. This is similar to other countries in sub-Saharan Africa (Faro et al., 2023; Mupara et al., 2022). Subsequently, delayed care-seeking and high relapse rates are common. Community-led support networks and improved linkage between hospital-based and community-based services are much needed in low-resource settings such as Kilimanjaro, Tanzania (Faro et al., 2023; Nadkarni et al., 2023; Turyasiima et al., 2025).

### Patient Experiences

All three participants expressed appreciation for and acceptability of the intervention, highlighting its pertinence. The participants praised the study nurses who improved their awareness and acceptance of the negative consequences of alcohol use, and for providing practical advice on how to reduce drinking. The qualitative feedback from the participants reinforces key interventionist traits that include empathy and non-judgmental listening. These elements are widely recognized as fundamental to effective counselling (Searight, 2007).

Remarkably, while the BI did not result in total cessation of drinking for all participants, the three cases emphasize the appreciation for the progress made regardless of objective change in scores over time. The participants recalled the valuable tools utilized for managing interpersonal challenges and the complex interplay of individual, social, and economic factors that influenced their drinking. This further illustrates the profundity and transformative potential of a brief and single interaction.

In all cases, participants valued the personalized nature of the MI approach particularly effective. The collaborative setting—where patients had the opportunity to express their concerns and goals—fostered a sense of agency over their behaviour change process (Smedslund et al., 2011). The negotiational approach to setting goals further increases ownership and acceptance of needed change, and enhances the integration of cognition and action (Radtchenko-Draillard, 2021). This finding underpins the importance of patient-centered care in addressing harmful alcohol use, particularly in settings where patients may not be familiar with counselling (Searight, 2007).

### Implications for Clinical Practice

This case series reveals that the use of a culturally adapted BI in the ED setting can be an effective means of reducing alcohol-related harms. This is particularly relevant in resource-limited settings, such as Tanzania, where mental health professionals are scarce, transitional support between acute care and recovery is lacking, and traditional treatment options for alcohol use disorder are also lacking (Knettel et al., 2023; Pauley et al., 2024; Staton et al., 2018).

The BI’s success in a busy ED highlights its feasibility and potential to be integrated into routine clinical practice without overwhelming existing healthcare resources. This argument is unavoidable in LMIC settings where cost-effectiveness, complexity of the intervention, and burden on resources determine the uptake of new interventions (Yamey, 2012).

The BI’s use of MI techniques are valuable in enhancing patient engagement and promoting behavior change. Allowing space and time for building rapport, exploring ambivalence, and fostering intrinsic motivation are crucial in addressing alcohol misuse (Smedslund et al., 2011). Training ED staff, including nurses, in MI techniques could increase the capacity of healthcare systems to address alcohol-related harms effectively (Searight, 2007). It is noteworthy that MI techniques are not exclusively effective for alcohol- or mental health-related conditions, but can be effectively employed by non-specialist staff to improve outcomes such as medication adherence, lifestyle change, and engagement in risk-taking behaviour which are all relevant to patients presenting at the ED (Institute of Medicine, 2003; Knettel et al., 2023).

Finally, the study emphasizes the need for a multi-disciplinary approach to alcohol-related interventions. While the BI provided a starting point for behaviour change, strengthened collaboration between ED and mental health settings is needed to achieve long-term success. Integrating mental health or counselling services into ED settings could be an important next step in enhancing the effectiveness of alcohol interventions (Staton et al., 2018, 2022).

### Implications for Policy

The success of the PPKAY intervention has important implications for public health policy in Tanzania and similar settings. Policymakers should strongly consider uptake and promotion of BIs into the standard care protocols for ED settings. The WHO’s SAFER initiative calls for the implementation of alcohol screening and brief interventions (World Health Organization, 2019), and this study exemplifies their potential success and relevance for LMICs, where resource limitations and the role of leadership must be acknowledged (Yamey, 2012).

Additionally, the current study recalls the importance of addressing broader socio-cultural factors that influence alcohol consumption in Tanzania, particularly the roles of alcohol in tradition and Tanzanians’ livelihood. Policy interventions should consider multisectoral strategies to reduce alcohol availability, particularly unregulated local brews, and promote public education on the harms of alcohol use (Ibitoye et al., 2019; Madundo et al., 2024).

Finally, the integration and promotion of mental health services should be prioritized in national health policies. This would ensure that individuals struggling with hazardous alcohol use have access to comprehensive care, which includes not only brief interventions but also long-term and transitional support (Mushi, Moshiro, et al., 2022).

### Implications for Future Research

Further studies are needed to explore the long-term effectiveness of BIs in reducing alcohol-related harms in Tanzania and other LMICs. Future research could assess whether the reductions in alcohol consumption and related harms are sustained beyond the 3-month follow-up period and whether additional interventions enhance the outcomes (Kaner et al., 2018).

Additional research is needed to examine the moderating and mediating factors that contribute to varying responses to the BI. Understanding the individual, socio-cultural, and economic factors that influence the success of BIs can help refine the intervention and tailor it more effectively to different settings and populations (Kaner et al., 2018; Platt et al., 2016). Research could also focus on who BIs do not benefit as a means to improve our understanding of failure to affect behaviour change (Edlind et al., 2018).

Future studies should explore the scalability of the PPKAY intervention in other healthcare settings and regions within Tanzania. Investigating the feasibility and effectiveness of PPKAY in settings with varying capacity, management, and infrastructure could provide valuable insights into how alcohol-related harms can be addressed across a broader range of contexts (Yamey, 2012).

## Limitations

Although we selected three participants representative of the PPKAY BI and its outcomes, our current methodology emphasizes in-depth analysis of individual cases, and our findings may not be generalizable to all participants or other settings.

This manuscript represents an exploration of the cases from the perspectives of the three patients. Participants may have felt compelled to emphasize the positive aspects of the BI and, although we conducted internal team supervision sessions to maximize accuracy of reporting and minimize bias, the potential for bias remains.

## Conclusion

This study illustrates that a culturally adapted BI can be an effective tool in reducing alcohol use and its associated harms in a Tanzanian ED setting. While the results demonstrate overall positive outcomes, the variability in participants’ responses underscores the complexity of hazardous alcohol use and the need for comprehensive, patient-centered, and multi-faceted interventions. We recommend integration of screening and BI for alcohol use in EDs, future research to enhance and explore the sustainability of intervention outcomes, and for policymakers to strongly consider promotion and integration of mental health services across Tanzania.

## Supporting information

CaRE Checklist

## Funding Statement

This project is supported by the National Institute on Alcohol Abuse and Alcoholism (NIAAA/NIH) (R01 AA027512). We also acknowledge the support from the NIH/Fogarty International Center D43 “The TRECK Program: Trauma Research Capacity Building in Kilimanjaro, Tanzania) (D43 TW012205) and NIAAA/NIH R34 “PRICE-Alcohol: Planning the Regional Implementation of a Culturally Adapted Brief Intervention for Alcohol for Tanzanian Emergency Departments” (R34AA031585).

## Author Contributions

KM contributed to conceptualization, methods, analysis, writing the first draft, and review and editing of the current study. AK contributed to methods, analysis, review and editing. AP led on study coordination, methods, review and editing. LM contributed to analysis, methods, review and editing. MP contributed to data collection, methods, and review and editing. JVPD contributed to coordination, data collection, methods, and review and editing. AA contributed to data collection, review and editing. JRNV contributed to analysis, review and editing. BM and CS contributed to initial trial design, trial funding and management, coordination, and review and editing.

## Data availability

Deidentified datasets used and/or analysed during the current study are available through URL upon request from Kennedy Ngowi at. KN is an ICT administrator, non-author, and ethics committee member at Kilimanjaro Clinical Research Institute.

## Notes

**Conflicts of interest:** The authors report no conflicts of interest.

### Competing Interest Statement

The authors have declared no competing interest.

### Clinical Trial

NCT04535011

### Author Declarations

Ethics committe/IRB of Duke University Health System (Pro000106116), Kilimanjaro Christian Medical Centre (Cert. #2486) and Tanzanias National Institute of Medical Research (HQ/R.8a/Vol.IX, 3425) gave ethical approval for this work.

