## Supplementary material for "The impact and experience of a culturally adapted brief intervention for harmful alcohol use in an emergency care setting (PPKAY: Punguza Pombe Kwa Afya Yako): A case series": CaRE Checklist

CARE case report guidelines

### CARE Checklist of information to include when writing a case report

(cc) BY-NC-ND

| Topic | Item | Checklist item description | Reported on Line |
|---|---|---|---|
| **Title** | 1 | The diagnosis or intervention of primary focus followed by the words "case report" | 1-2 |
| **Keywords** | 2 | 2 to 5 key words that identify diagnoses or interventions in this case report, including "case report" | 64 |
| **Abstract** | 3a | Introduction: What is unique about this case and what does it add to the scientific literature? | 36-40 |
| | 3b | Main symptoms and/or important clinical findings | 41-51 |
| | 3c | The main diagnoses, therapeutic interventions, and outcomes | 42-57 |
| | 3d | Conclusion—What is the main "take-away" lesson(s) from this case? | 58-62 |
| **Introduction** | 4 | One or two paragraphs summarizing why this case is unique (**may include references**) | 105-108 |
| **Patient information** | 5a | De-identified patient specific information | 229-233; 278-281; 326-328 |
| | 5b | Primary concerns and symptoms of the patient | 234-241; 282-289; 329-336 |
| | 5c | Medical, family, and psycho-social history including relevant genetic information | 234-244; 282-292; 329-339 |
| | 5d | Relevant past interventions with outcomes | Not applicable |
| **Clinical findings** | 6 | Describe significant physical examination (PE) and important clinical findings | 234-241; 282-289; 329-336 |
| **Timeline** | 7 | Historical and current information from this episode of care organized as a timeline | 234-259; 282-316; 329-350 |
| **Diagnostic information** | 8a | Diagnostic testing (such as PE, laboratory testing, imaging, surveys) | 222-223; 260; 322-333 |
| | 8b | Diagnostic challenges (such as access to testing, financial, or cultural) | 235-236; 284-285; 331-332 |
| | 8c | Diagnosis (including other diagnoses considered) | Not applicable |
| | 8d | Prognosis (such as staging in oncology) where applicable | 261-264; 317-323; 351-354 |
| **Intervention** | 9a | Types of therapeutic intervention (such as pharmacologic, surgical, preventive, self-care) | 257-259; 314-316; 348-350 |
| | 9b | Administration of therapeutic intervention (such as dosage, strength, duration) | 158-168 |
| | 9c | Changes in therapeutic intervention (with rationale) | 143-144 |
| **Outcomes** | 10a | Clinician and patient-assessed outcomes (if available) | 261-264; 317-323; 351-354 |
| | 10b | Important follow-up diagnostic and other test results | 261-264; 317-323; 351-354 |
| | 10c | Intervention adherence and tolerability (How was this assessed?) | 189-192 |
| | 10d | Adverse and unanticipated events | 193 |
| **Discussion** | 11a | A scientific discussion of the strengths AND limitations associated with this case report | 375-531 |
| | 11b | Discussion of the relevant medical literature **with references** | 375-523 |
| | 11c | The scientific rationale for any conclusions (including assessment of possible causes) | 462-523 |
| | 11d | The primary "take-away" lessons of this case report (without references) in a one paragraph conclusion | 533-540 |
| **Patient perspective** | 12 | The patient should share their perspective in one to two paragraphs on the treatment(s) they received | 275-276; 306-308; 355-357 |
| **Informed consent** | 13 | Did the patient give informed consent? Please provide if requested | **Yes** ☑ **No** ☐ |
